# CDH26 amplifies airway epithelial IL-4 receptor α signaling in asthma

**DOI:** 10.1101/2020.12.01.20241752

**Authors:** Yuchen Feng, Shengchong Chen, Chenli Chang, Wenliang Wu, Dian Chen, Jiali Gao, Gongqi Chen, Lingling Yi, Guohua Zhen

## Abstract

**Background:** Activation of interleukin (IL)-4 receptor (R) signaling in airway epithelial cells leads to airway hyperresponsiveness and mucus overproduction in asthma. Cadherin-26 (CDH26), a cadherin implicated in polarization of airway epithelial cells, is upregulated in asthma. However, the role of CDH26 in asthma remains unknown. We hypothesize that CDH26 plays a role in airway epithelial IL-4R signaling in asthma.

**Methods:** We measured airway resistance, mucus production, airway inflammation, and Il-4Rα expression in *Cdh26*^-/-^ and WT mice after allergen sensitization and challenge. We explored the role of CDH26 in IL-4R signaling, mucin genes and eosinophilic chemokine expression in cultured bronchial epithelial cells and bronchial brushings from asthma patients.

**Results:** Cdh26 deficiency nearly blocked airway mucus overproduction, and suppressed AHR and airway eosinophilia in a murine model of allergic airway disease. Interestingly, Il-4Rα expression in airway epithelium was markedly reduced in *Cdh26*^*-/-*^ mice. In cultured human bronchial epithelial cells, CDH26 knockdown inhibited IL-13, a ligand for IL-4R, -induced IL-4Rα and IL-13Rα1 expression, and suppressed the downstream Jak1 and Stat6 phosphorylation. Moreover, CDH26 knockdown inhibited IL-13-induced MUC5AC, MUC5B and eosinophilic chemokines CCL11, CCL24, CCL26 expression. In contrast, CDH26 overexpression intensified IL-13-induced activation of IL-4Rα signaling. In asthma patients, CDH26 was the only one upregulated of 11 cadherins in bronchial brushings. CDH26 expression significantly correlated with epithelial IL-4Rα, MUC5AC expression, sputum eosinophilia and fractional exhaled nitric oxide (FeNO).

**Conclusion:** Taken together, CDH26 is an amplifier of epithelial IL-4R signaling in asthma, and may represent a therapeutic target for airway mucus overproduction.

## INTRODUCTION

Asthma is a chronic inflammatory airway disease affecting more than 300 million people worldwide^1,2^. Asthma has different phenotypes and endotypes, and type 2-high asthma is a prominent endotype characterized by airway hyperresponsiveness (AHR), airway mucus overproduction and eosinophilia^3,4^. Type 2 cytokines including interleukin (IL)-4 and IL-13 play critical roles in the pathogenesis of type 2-high asthma. Both of IL-4 and IL-13 activate IL-4 receptor (R) signaling in hematopoietic immune cells and non-hematopoietic structural cells including airway smooth muscle cells and epithelial cells, leading to AHR, airway eosinophilia and mucus overproduction in asthma^5–9^.

IL-4R signaling pathway play a pivotal role in the pathogenesis of asthma and other allergic diseases such as helminth infection and atopic dermatitis^6,10–12^. Type I IL-4R consisted of IL-4Rα and common γ chain binds to IL-4 exclusively, and is predominantly expressed by hematopoietic cells. IL-4Rα subunit pairs with IL-13Rα1 to form type II IL-4R which can bind to both IL-4 and IL-13 and is mainly expressed in non-hematopoietic cells including epithelial cells^13–15^. The downstream cascades of IL-4R signaling include phosphorylation of Janus kinase 1 (Jak1) and signal transducer and activator of transcription 6 (Stat6)^6,13,15^. Airway mucus cell metaplasia and mucus overproduction are strictly dependent on the activation of IL-4R signaling in airway epithelial cells^16^. *Stat6*^*-/-*^ mice failed to develop allergen induced AHR and mucus overproduction^5^. However, reconstitution of Stat6 only in epithelial cells of *Stat6*^*-/-*^ mice was sufficient for IL-13-induced AHR and mucus production^17^. Moreover, IL-4R signaling also mediates the production of eosinophilic chemokines including CCL11/eotaxin-1, CCL24/eotaxin-2 and CCL26/eotaxin-3 by airway epithelial cells to regulate airway eosinophilia^18–20^. Several studies reported potential mechanisms underlying IL-4R upregulation in immune cells and IL-4R degradation^21,22^. IL-4Rα expression was reported to be upregulated in nasal epithelial cells from asthmatic patients^21^. However, the mechanism by which the epithelial IL-4R is upregulated remains unknown.

Cadherins are a superfamily of transmembrane proteins which mediate calcium-dependent cell-cell adhesion and exhibit tissue-specific expression patterns^23^. Cadherin-26 (CDH26) is reported to be expressed in gastric and airway epithelial cells, and is an integrin-binding epithelial receptor implicated in allergic gastrointestinal inflammation^24–27^. Moreover, CDH26 plays a role in apicobasal polarization of airway epithelial cells^27^. Type 2 cytokine such as IL-13 induces CDH26 expression in human bronchial epithelial cells^24^. CDH26 upregulation has been reported in diseases characterized by type 2 inflammation including asthma, helminth infection, eosinophilic gastritis and esophagitis^25,26,28–31^. Although elevated epithelial CDH26 expression has been linked to asthma^31,32^, the role of CDH26 in asthma remains unknown. We hypothesize that CDH26 regulates airway epithelial IL-4R signaling in asthma.

In this study, we demonstrated that *Cdh26*^-/-^ mice exhibited reduced epithelial Il-4Rα expression, alleviated AHR, mucus overproduction and eosinophilia after allergen challenge. CDH26 upregulates IL-4Rα expression and amplifies the IL-4R signaling pathway, and promotes IL-13-induced mucin gene expression and eosinophilic chemokines expression in airway epithelial cells. In human asthma, CDH26 correlates with enhanced IL-4Rα, MUC5AC expression and parameters reflecting airway eosinophilia.

## MATERIALS AND METHODS

### Subjects

We recruited 17 healthy control subjects and 56 asthma patients. All subjects were Chinese and recruited from Tongji Hospital. Subject characteristics are summarized in Table S1. Asthma patients had symptoms of recurrent episodes of wheezing, breathlessness, chest tightness, and coughing, and had accumulated dosage of methacholine provoking a 20% fall (PD20) of forced expiratory volume in the first second (FEV1) <2.505 mg and/or ≥12% increase in FEV1 following inhalation of 200 μg salbutamol. Healthy control subjects had no respiratory symptoms, normal spirometry value, and methacholine PD20 ≥ 2.505 mg. None of the subjects had ever smoked or received inhaled or oral corticosteroid or leukotriene antagonists. For each subject, we recorded demographic information, collected induced sputum, and measured spirometry and FeNO. We performed bronchoscopy with endobronchial brushing and biopsy. Biopsy techniques and methods for spirometry and FeNO measurement were described previously^33^. Written informed consent was obtained from all subjects. The ethics committee of Tongji Hospital, Huazhong University of Science and Technology, approved the study.

### Animals

*Cdh26*^*-/-*^ mice on a C57BL/6 background were purchased from the KOMP Repository Collection (UC Davis, USA), and exon 2 to exon 12 of Cdh26 gene was knocked out. Wild-type (WT) C57BL/6 mice were obtained from the Animal Experimental Center of Hubei Province (Wuhan, China). All experimental procedures were approved by the Animal Care and Use Committee of the Tongji Hospital, Huazhong University of Science and Technology.

### Murine model of allergic airway disease

*Cdh26* ^*-/-*^mice and WT mice received 30 μg HDM in 40ul (Greer Laboratories, Lenoir, NC, USA) or saline (controls) intraperitoneally, once a week for three weeks before receiving three challenges for three days with 125 μg HDM in 40 ul or the same volume of saline, intranasally. Twenty-four hours after the last challenge, we measured pulmonary resistance in response to a range of intravenous methacholine concentrations using the forced oscillation technique with the FlexiVent system (SCIREQ). Then bronchoalveolar lavage fluid (BALF) was collected, and inflammatory cells were collected by low-speed centrifugation. BALF cell counts for macrophages, eosinophils, lymphocytes, and neutrophils in bronchoalveolar lavage fluid were performed as previously described^34^. Lung tissues were collected for histological analysis, quantitative PCR, and immunostaining.

### Cell culture and treatment

Human bronchial epithelial (HBE) cells collected from healthy donors (n = 3) by bronchial brushing technique were cultured at an air-liquid interface as previously described^35,36^. BEAS-2B cell lines were purchased from ATCC (Manassas, VA). Cells were cultured in DMEM medium with 10% FBS (Biological Industries, Kibbutz Beit-Haemek, Israel) and were transfected with scrambled control or CDH26 siRNA (80 nM; GeneCopoeia), or empty or CDH26 cDNA expression vector (500 ng/mL) using Lipofectamine 3000 (Invitrogen, Carlsbad, CA). Forty-eight hours before the end of culture, IL-13 (20 ng/mL; PeproTech) was added to the basal medium. Finally, cells were harvested for quantitative PCR and Western blotting. The cell culture media were collected for ELISA. The sequence of CDH26 siRNA are described in Supplementary Materials.

### Quantitative PCR

Total RNA from bronchial epithelial brushing, mouse lungs, HBE cells, and BEAS-2B cells was isolated using TRIzol (Invitrogen, Carlsbad, CA) and reverse transcribed to cDNA using PrimeScript RT reagent kit (Takara, Tokyo, Japan). The sequences of the primers for Sybr Green real-time PCR were obtained from PrimerBank. Other primer sequences were designed by Sangon Biotech (Sangon Biotech, SH, China). The transcript levels of *CDH26, CCL11, CCL24, CCL26, MUC5AC, MUC5B IL-4Rα, IL-13, IL-4*, and Cadherin family members were determined by using Takara SYBR Premix ExTaq polymerase and a CFX Connect PCR System (Bio-Rad Laboratories, California, USA) Fold differences were determined by the 2-ΔΔCT method^37^. The transcript levels of each gene are expressed as relative to the median of healthy control subjects or the mean of the control group, and log2 transformed. Primers for quantitative PCR were listed in the Supplementary Table S2.

### Histology and immunofluorescence staining

The mouse left lungs were inflation-fixed and paraffin-embedded, and 5-μm sections were cut. Sections were used for hematoxylin and eosin staining, PAS staining, and immunohistochemical examination. Colocalization of Cdh26 and Il-4Rα were analyzed using ImageJ, Using the ImageJ plug-in Coloc2, Pearson’s correlation coefficient and Manders’ coefficient were calculated to assess colocalization of Cdh26 and Il-4Rα as previously described^38^. The methods for immunohistochemistry, assessment of airway inflammation and PAS staining are described in Supplementary Materials.

### Western blotting

Proteins were extracted from cells or lung tissues using RIPA buffer (Servicebio, Wuhan, China). Fifty micrograms of extracted proteins were separated using 10% SDS-PAGE, and the separated proteins were transferred onto polyvinylidene difluoride (PVDF) membranes (Roche, Mannheim, Germany). The membranes were first probed with indicated primary antibodies. Antibodies used in western blot were:CDH26 (Sigma-Aldrich, 1:1000 dilution), mouse IL-4Rα antibody (R&D system, 1:2000 dilution), human IL-4Rα antibody (R&D system, 1:500 dilution), total and phosphorylated Stat6 (Cell Signaling, 1:1,000 dilution), total and phosphorylated Jak1 (Cell Signaling, 1:1000 dilution), GAPDH (Aspen, 1:2000 dilution). Then antibody was detected using horseradish peroxidase-conjugated goat anti-rabbit IgG (Aspen, 1:4000 dilution) or goat anti-mouse IgG (Aspen, 1:4000 dilution) or rabbit anti-goat IgG secondary antibody followed by ECL Western blot detection reagent (Beyotime Biotech). Densitometry was assessed using ImageJ (National Institutes of Health) and calculated according to the reference bands of GAPDH.

### ELISA

CCL11, CCL24, and CCL26 in BEAS-2B cell culture media were analyzed by ELISA (R&D systems, Bio-Techne, USA). And Ccl11, Ccl24 in BALF were also analyzed by ELISA (R&D systems, Bio-Techne, USA). ELISA was performed according to the manufacturer’s instructions. All samples and standards were measured in duplicate.

### Statistical analysis

Distributed data are presented as means ± standard deviation (SD), and P-values were calculated using parametric tests (Student’s t-test or one-way ANOVA followed by Tukey’s multiple comparison test). For non-normally-distributed data, we calculated medians with interquartile ranges and used non-parametric tests (Kruskal-Wallis test with Dunn intergroup comparison or Mann-Whitney test). We analyzed correlation using Spearman’s rank-order correlation. Statistical analysis was performed with GraphPad Prism version 7.1 (GraphPad Software, San Diego, CA, USA). Statistical significance was established at P < 0.05.

## RESULTS

### Cdh26 deficiency suppresses AHR and nearly blocks mucus overproduction

To explore the role of CDH26 in asthma, *Cdh26*^-/-^ mice were used to establish a model of allergic airway disease (Figure S1). Wild type (WT) and *Cdh26*^-/-^ mice were sensitized and challenged with house dust mite (HDM) (Figure 1A). HDM challenge significantly increased Cdh26 protein expression in mice lung tissue as analyzed by Western blotting (Figure 1B). Immunostaining revealed that Cdh26 protein expression was localized in airway epithelial cells, and the fluorescent intensity of Cdh26 in the airway was markedly increased after the HDM challenge (Figure S1B).

**Figure 1.**
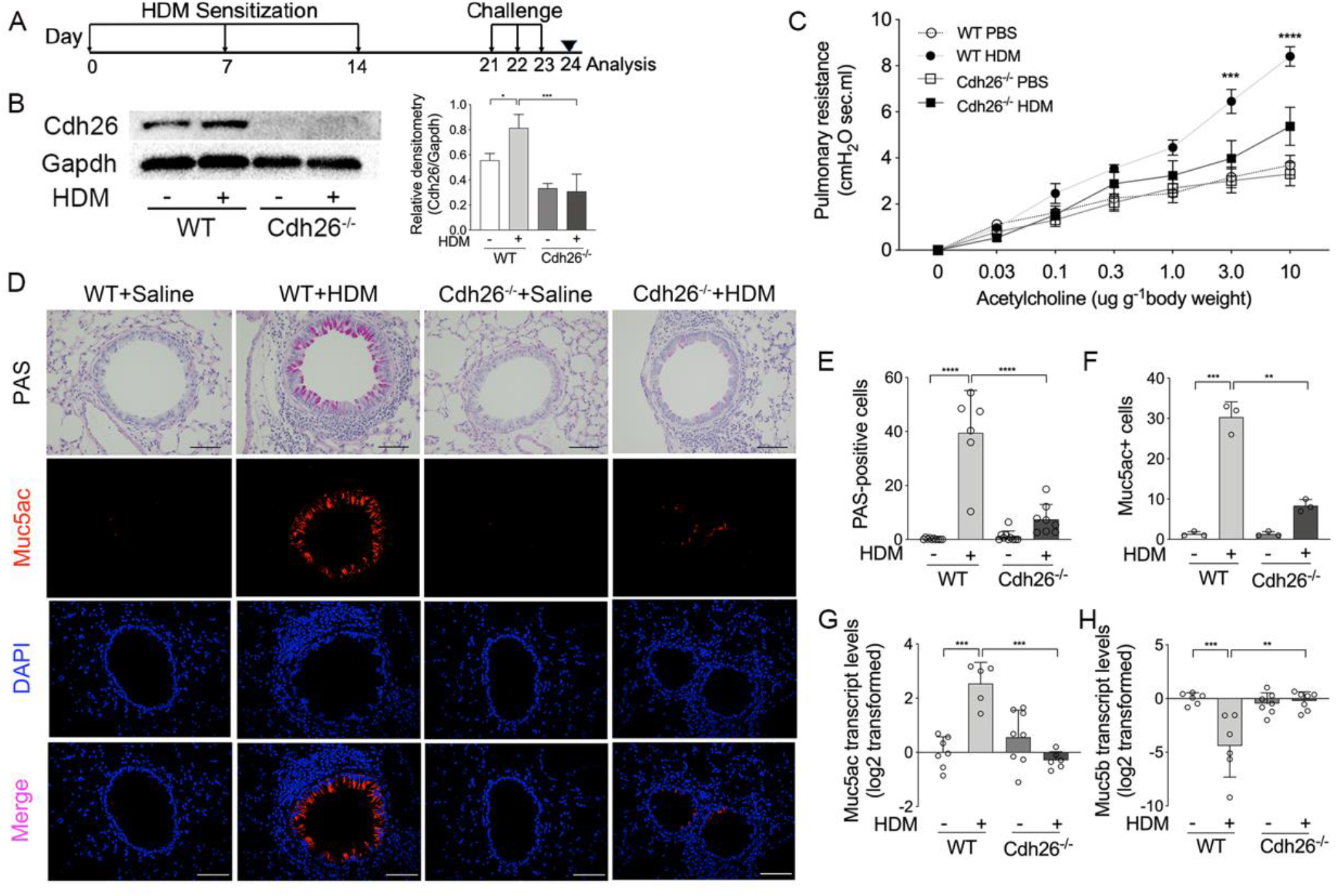
CDH26 deficiency suppresses AHR and nearly blocks mucus overproduction. **A)** Experimental schedule. **B)** Cdh26 protein levels in mouse lungs were measured by Western blotting. Densitometry assay was performed using ImageJ, and Cdh26 protein levels were indexed to Gapdh. **C)** Pulmonary resistance in response to different concentration of intravenous methacholine in WT mice and *Cdh26*^*-/-*^ mice that were administered intranasally with HDM or saline. **D)** Representative images of PAS staining, Muc5ac immunofluorescence staining and DAPI staining in mouse lung sections. Scale bar, 50 μm. **E)** The numbers of PAS-staining-positive cells were counted in four random fields for each lung section at 200× magnification. **F)** The numbers of Muc5ac-staining-positive cells were counted in five random fields for each lung section at 200× magnification. **G, H)** The mRNA levels of *Muc5ac* (*G*) and *Muc5b* (*H*) in mouse lung were determined by quantitative PCR. The transcript level was expressed as log2 transformed and relative to the mean of control group. n =5-8 mice per group. Data are mean ± SD. *P<0.05; **P<0.01; ***P<0.001; ****P < 0.0001 (one-way ANOVA followed by Tukey’s multiple comparison test). The data are representative of three independent experiments.

We first examined the role of Cdh26 in airway hyperresponsiveness (AHR) and mucus overproduction. In WT mice, the airway resistance to methacholine was increased after HDM sensitization and challenge, and the number of PAS staining-positive mucus cells and the expression of Muc5ac were also markedly enhanced (Figure 1C-1F). However, HDM-induced increase of airway resistance was significantly suppressed in *Cdh26*^-/-^ mice compared to WT mice (Figure 1C). Remarkably, HDM-induced mucus cell hyperplasia and Muc5ac upregulation were nearly blocked in *Cdh26*^-/-^ mice when compared with WT mice. PAS-positive cells were reduced by 86% and Mac5ac-positive cells were reduced by 75% in HDM-challenged *Cdh26*^-/-^ mice compared to WT mice. Using quantitative PCR, we found that HDM challenge increased *Muc5ac* transcripts and decreased *Muc5b* transcripts in lungs of WT mice, both of which were completely suppressed in *Cdh26*^-/-^ mice (Figure 1G, H). Our data suggest that upregulated airway epithelial Cdh26 promotes AHR and mucus overproduction in asthma.

### Cdh26 deficiency suppresses lung eosinophilic inflammation

We next explored the role of Cdh26 in the allergic airway inflammation of the mouse model. HDM sensitization and challenge led to peri-bronchial eosinophilic inflammation in WT mice as shown by HE staining (Figure 2A). Using inflammatory scores to quantify the extent of the inflammation, HDM-induced inflammation was slightly alleviated in *Cdh26*^*-/-*^ mice compared to WT mice (Figure 2B). Meanwhile, the HDM challenge increased the number of total cells and eosinophils in WT mice’s airways as analyzed by BALF cell counting and differentiation (Figure 2C). However, HDM-induced increase of total cells and eosinophils were significantly reduced in *Cdh26*^*-/-*^ mice (Figure 3). Moreover, the expression of eosinophilic chemokines Ccl11, Ccl24, Ccl26 were increased in BALF supernatant or lung tissue from HDM-challenged WT mice, but the HDM-induced expression of these chemokines was suppressed in *Cdh26*^*-/-*^ mice (Figure 2D-2F). Our data suggest that Cdh26 contributes to airway eosinophilic inflammation, at least in part, via promoting eosinophilic chemokines expression.

**Figure 2.**
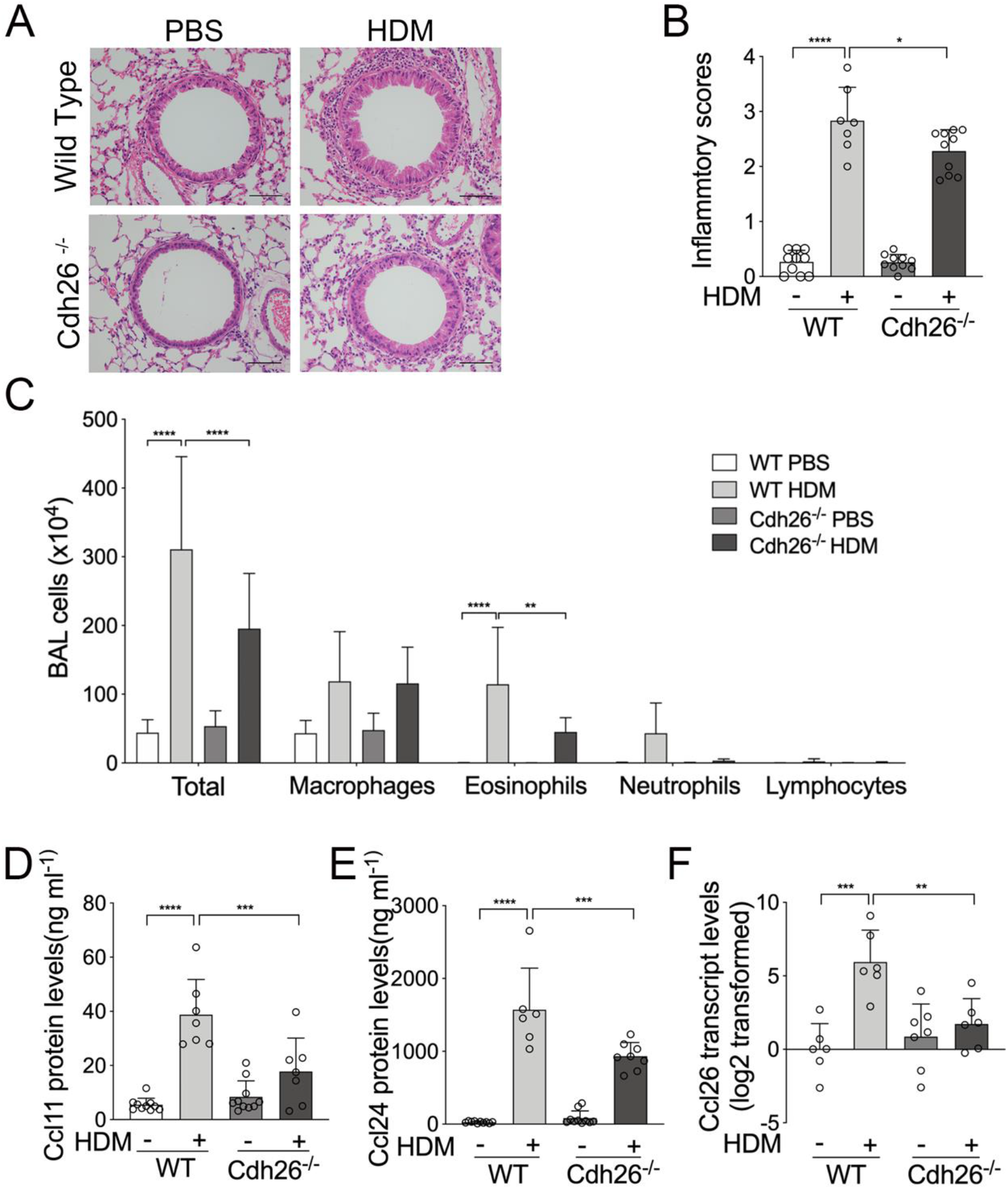
Cdh26 deficiency suppresses lung inflammation. **A)** Representative images of H&E staining of mouse lung sections. Scale bar, 50 μm. **B)** Lung inflammatory scores were calculated as described in Methods. **C)** Cell counting for macrophages, eosinophils, lymphocytes and neutrophils in BALF. **D, E)** The protein levels of Ccl11(*D*), Ccl24(*E*) in BALF were determined by ELISA. **F)** The mRNA levels of Ccl26 in mouse lungs were determined by quantitative PCR. The transcript level was expressed as log2 transformed and relative to the mean of control group. n =5-8 mice per group. Data are mean ± SD. *P<0.05; **P<0.01; ***P<0.001; ****P < 0.0001 (one-way ANOVA followed by Tukey’s multiple comparison test). The data are representative of three independent experiments.

**Figure 3.**
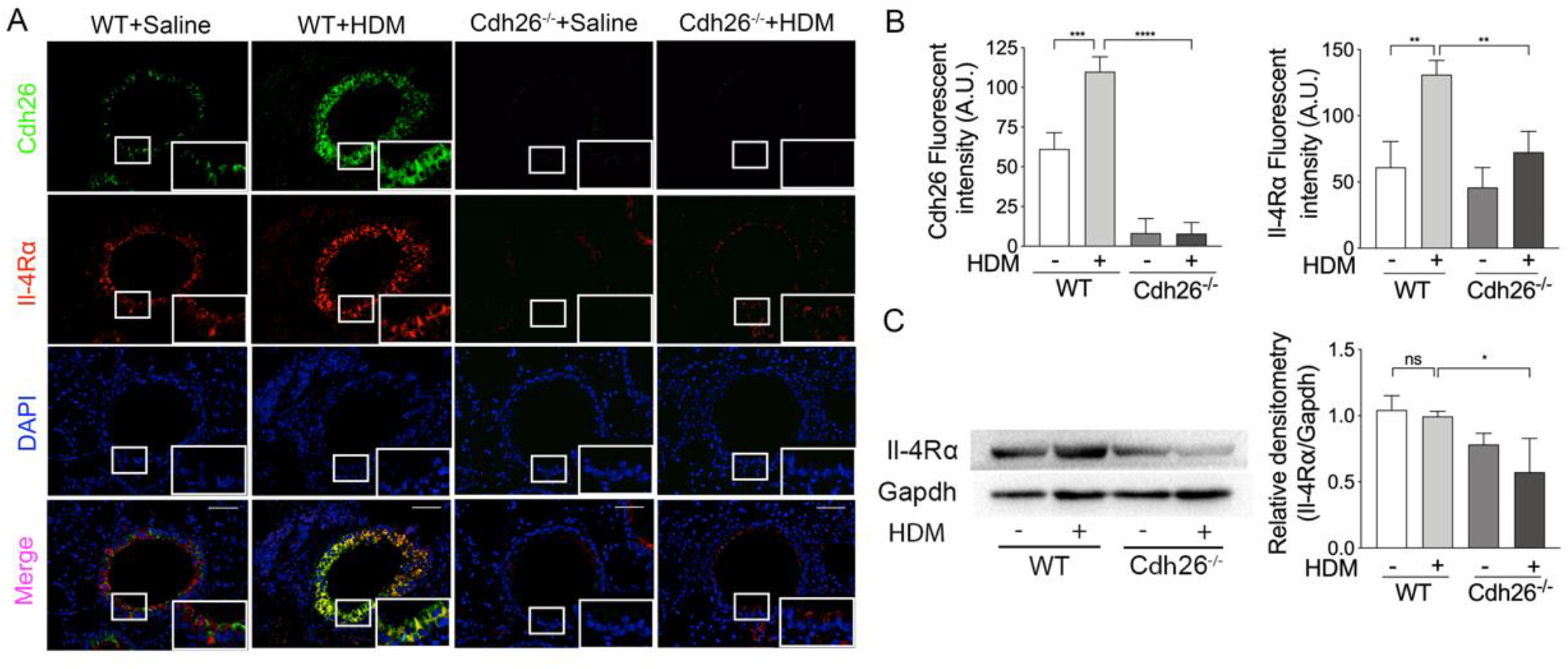
Cdh26 upregulates airway epithelial Il-4Rα expression in mice. **A)** Representative images of Cdh26 (green) and Il-4Rα (red) immunofluorescence staining in mouse lung sections. Nuclei were stained with DAPI (blue). Scale bar, 50 μm. **B)** Quantification of Cdh26 and Il-4Ra immunofluorescent staining of WT and *Cdh26*^*-/-*^ mice treated with Saline or HDM. Fluorescent intensity assay was performed using ImageJ. **C)** Il-4Rα protein levels in mouse lungs were measured by Western blotting. Densitometry assay were performed using ImageJ, and the protein levels of Il-4Rα were indexed to Gapdh. Data are mean ± SD. *P<0.05; **P<0.01; ***P<0.001; ****P < 0.0001 (one-way ANOVA followed by Tukey’s multiple comparison test). The data are representative of three independent experiments.

### Cdh26 upregulates airway epithelial Il-4Rα expression in mice

Since airway mucus cell hyperplasia and aberrant expression of mucin genes are strictly dependent on IL-4R signaling in airway epithelial cells^17,39^, we next examined whether Cdh26 deficiency affects airway epithelial Il-4Rα expression. Immunostaining showed that Il-4Rα was expressed in the airway epithelium of WT mice, and the HDM challenge significantly increased the fluorescent intensity of Il-4Rα (Figure 3A). However, quantification of immunostaining in airway epithelium showed that HDM-induced epithelial Il-4Rα expression was markedly reduced in *Cdh26*^*-/-*^ mice compared to WT mice. Actually, the fluorescent intensity of epithelial Il-4Rα expression in HDM-challenged *Cdh26*^*-/-*^ mice was almost similar to that in PBS-challenged WT and *Cdh26*^*-/-*^ mice (Figure 3B). This indicates that Cdh26 is required for allergen-induced epithelial Il-4Rα upregulation in allergic airway disease. Moreover, Il-4Rα protein levels in the whole lung tissue were also significantly reduced in *Cdh26*^*-/-*^ mice compared to WT mice after HDM challenge as analyzed by Western blotting (Figure 3C). However, the HDM challenge did not increase Il-4Rα protein levels in whole lung tissue, probably because the upregulation of Il-4Rα was dominant in airway epithelium which can not be distinguished in the whole lung tissue. Our data suggest that Cdh26 upregulates epithelial Il-4Rα expression in asthma. In addition, immunostaining showed that Cdh26 colocalized with Il-4Rα in mouse airway epithelium (Figure 3A). When we measured the fluorescent pixels positive for Cdh26 or Il-4Rα and analyzed with Pearson’s and Manders’ correlation coefficient, the pixels positive for both Cdh26 and Il-4Rα accounted for 80% of the pixels positive for Cdh26 or Il-4Rα, respectively (Figure S2).

### CDH26 amplifies IL-4R—Jak—Stat6 pathway in airway epithelial cells

Type 2 IL-4R consisting of IL-4Rα and IL-13Rα1 is predominant in airway epithelial cells ^14,40^. In cultured human bronchial epithelial (HBE) cells, we further investigated the role of CDH26 in the expression of IL-4Rα and IL-13Rα1 and the downstream signaling cascades including Jak1 and Stat6 phosphorylation. We found that IL-13 increased *CDH26* mRNA expression in HBE cells cultured on air-liquid interface (ALI) at 12, 24, 48h (Figure 4A). CDH26 protein expression was also increased after IL-13 treatment for 48h in human BEAS-2B bronchial epithelial cell line (Figure 4B). IL-13 stimulation increased IL-13Rα1 expression and tended to increase IL-4Rα expression in BEAS-2B cells (Figure 4C). Importantly, IL-13-induced IL-4Rα and IL-13Rα1 expression were significantly suppressed when CDH26 gene expression was knockdown. Further, Jak1 and Stat6 phosphorylation were markedly increased after IL-13 treatment for 0.5h in BEAS-2B cells. However, IL-13-induced Jak1 and Stat6 phosphorylation were dramatically reduced when CDH26 expression was knockdown (Figure 4E). Interestingly, CDH26 overexpression further enhanced IL-13-induced IL-4Rα and IL-13Rα1 expression as well as Jak1 and Stat6 phosphorylation (Figure S3). Our data indicate that IL-13, a IL-4R ligand, upregulates CDH26 expression, which in turn amplifies the activation of IL-4R-Jak1-Stat6 signaling in airway epithelial cells, forming a feed-forward loop.

**Figure 4.**
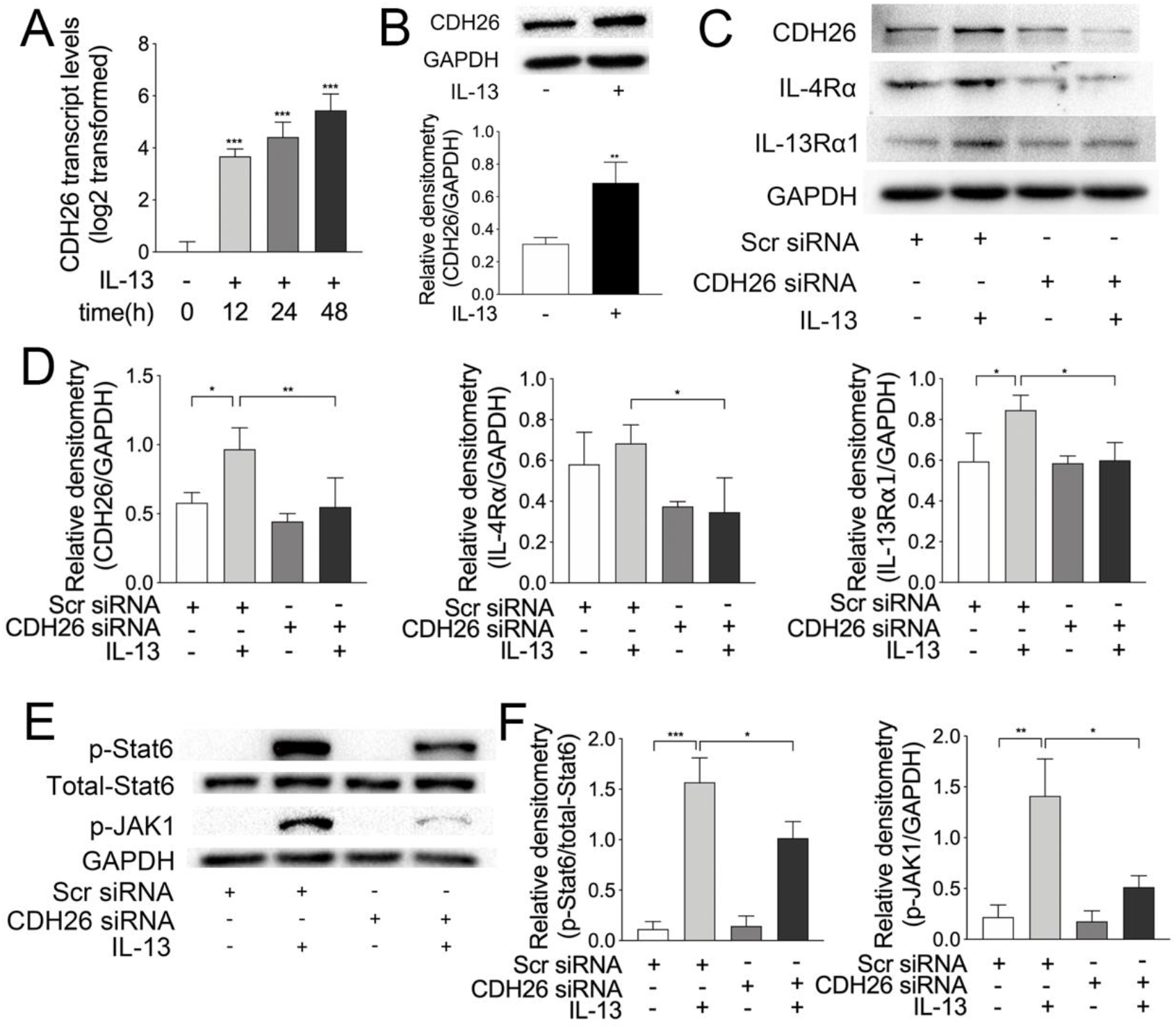
CDH26 amplifies IL-4R—Jak1—Stat6 pathway in airway epithelial cells. **A)** HBE cells were cultured at air-liquid interface for 14 d and was then stimulated with IL-13 for 12h, 24h and 48h. The mRNA levels of *CDH26* in HBE cells were determined by quantitative PCR. n=3 wells per group. The transcript level was expressed as log2 transformed and relative to the mean of control group. **B)** The protein levels of CDH26 in BEAS-2B cells after exposure to IL-13 for 48h were determined by Western blotting. Densitometry assay was performed using ImageJ, and CDH26 protein levels were indexed to GAPDH. The data are representative of three independent experiments. **C)** The protein levels of IL-4Rα, IL-13Rα1 in BEAS-2B cells after transfection with control or CDH26 siRNA with or without IL-13 treatment for 48h were detected by Western blotting. **D)** Densitometry assay of the Western blotting results was performed using ImageJ. The protein levels of CDH26, IL-4Rα and IL-13Rα1 were indexed to GAPDH. **E)** The protein levels of phosphorylated and total Stat6, phosphorylated Jak1 in BEAS-2B cells after transfection with control or CDH26 siRNA with or without IL-13 treatment for 30min were detected by Western blotting. **F)** Densitometry assay of phosphorylated Jak1 relative to total GAPDH and phosphorylated Stat6 relative to total STAT6. The data are representative of three independent experiments. Data are mean ± SD. *P<0.05; **P<0.01; ***P<0.001 (one-way ANOVA followed by Tukey’s multiple comparison test).

### CDH26 promotes MUC5AC and eosinophilic chemokines expression in airway epithelial cells

Given the amplifying role of CDH26 amplifies in epithelial IL-4R signaling, we asked whether CDH26 knockdown alters IL-13-induced mucin gene expression in HBE cells. IL-13 treatment for 48 h upregulated *MUC5AC* mRNA expression and downregulated *MUC5B* mRNA expression in HBE cells cultured on ALI. Consistent with our findings in HDM-challenged *Cdh26*^*-/-*^ mice, CDH26 knockdown significantly suppressed IL-13-induced *MUC5AC* upregulation and *MUC5B* downregulation (Figure 5A-5B).

**Figure 5.**
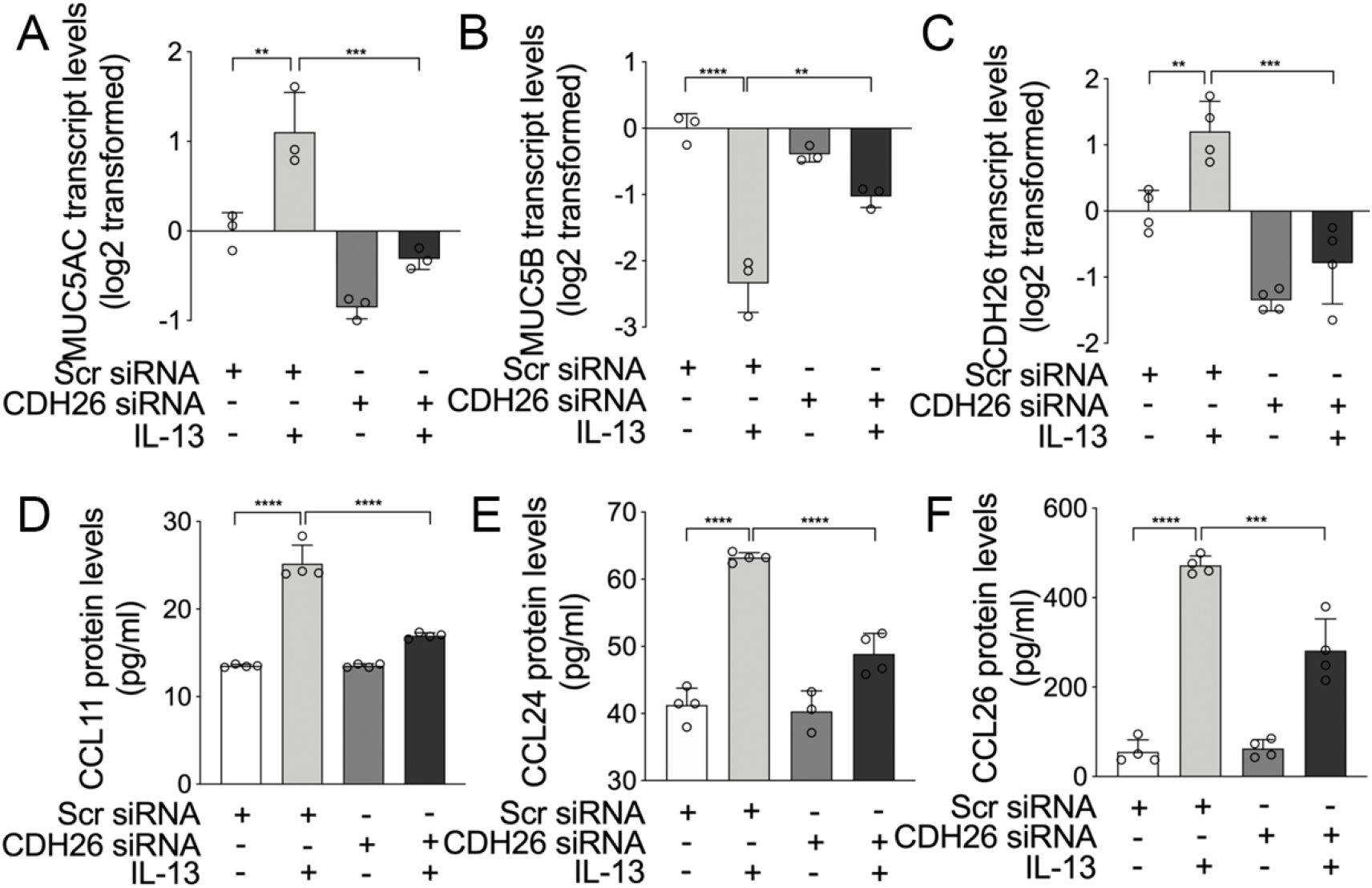
CDH26 promotes MUC5AC and eosinophilic chemokines expression in airway epithelial cells. **A, B)** Primary culture of HBE cells at ALI for 14 d with IL-13 stimulation for 72h of culture. The transcript levels of *MUC5AC* (*A*) and *MUC5B* (*B*) in HBE cells after transfection with control or CDH26 siRNA with or without IL-13 stimulation were determined by quantitative PCR. The transcript levels were expressed as log2 transformed and relative to the mean of control group. **C)** The transcript levels of *CDH26* in BEAS-2B cells after transfection with control or CDH26 siRNA with or without IL-13 stimulation for 48h were determined by quantitative PCR. n = 4 wells per group. **D-F)** The protein levels of CCL11 (*D*), CCL24 (*E*) and CCL26 (*F*) in the culture media of BEAS-2B cells after transfection with control or CDH26 siRNA with or without IL-13 stimulation were detected by ELISA. n = 4 wells per group. Data are mean ± SD. *P < 0.05; **P < 0.01; ***P < 0.001; ****P < 0.0001 (one-way ANOVA followed by Tukey’s multiple comparison test). The data are representative of four independent experiments.

IL-4R signaling pathway was reported to contribute to the expression of eosinophilic chemokines CCL11, CCL24, CCL26^7,18,41^. Consistent with these reports, we found that Jak inhibitor Ruxolitinib blocked IL-13-induced *CCL11, CCL24*, and *CCL26* mRNA expression in BEAS-2B cells (Figure S4). IL-13 treatment increased the protein levels of these chemokines in supernatant of cell cultures, whereas CDH26 knockdown markedly suppressed IL-13-induced CCL11, CCL24 and CCL26 production (Figure 5C-5F). In contrast, CDH26 overexpression further enhanced IL-13-induced expression of these chemokines (Figure S5). Our data suggest that CDH26 promotes IL-4R signaling-dependent mucin gene expression and eosinophilic chemokines expression in airway epithelial cells.

### Epithelial CDH26 expression correlates with IL-4Rα expression, mucin gene expression, and airway eosinophilia in asthma

Finally, we examined CDH26 and IL-4Rα expression in asthma patients and healthy controls. *CDH26* transcript levels in bronchial brushings were markedly increased in asthma patients compared to healthy controls (Figure 6A). Moreover, *CDH26* was the only upregulated cadherin of the 11 cadherin family members in bronchial brushings from asthma patients (Figure 6B). *IL-4Rα* transcript levels were also increased in bronchial brushings from asthma patients (Figure 6C). Consistent with our findings in mice and cultured airway epithelial cells, there was a positive correlation between *CDH26* and *IL-4Rα* transcript levels (Figure 6D). Moreover, both *CDH26* and *IL-4Rα* transcript levels significantly correlate with 3-gene-mean, the mean transcript levels of IL-13-responsive epithelial genes *CLCA1, SERPINB2*, and *POSTN* (Figure 6E-6F). This suggests that epithelial *CDH26* and *IL-4Rα* expression associate with the type 2 status of asthma. Similarly, CDH26 and IL-4Rα-positive staining in bronchial biopsies were markedly increased in asthma patients compared to controls (Figure 6G-6H). Consistent with our finding in mice, CDH26 and IL-4Rα protein colocalized with each other in bronchial epithelial cells (Figure 6G).

**Figure 6.**
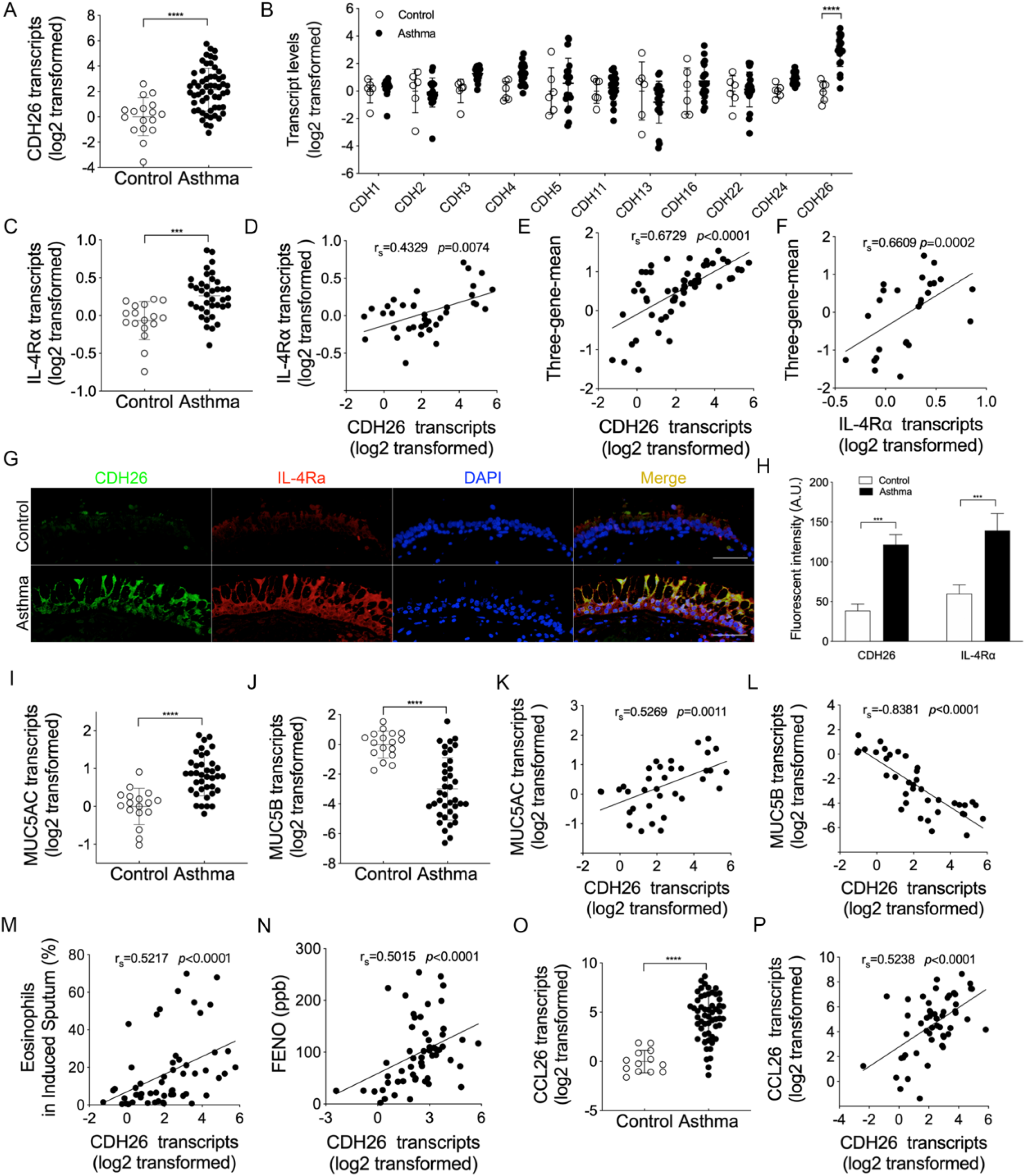
Epithelial CDH26 expression correlates with IL-4Rα expression, mucin gene expression, and airway eosinophilia in asthma. **A)** *CDH26* transcript levels in bronchial brushings from asthma patients (n=56) and healthy controls (n = 17) were measured by quantitative PCR. The transcript levels were expressed as log2 transformed and relative to the median value for healthy controls (two-tailed Mann– Whitney test). **B)** The transcript levels of cadherin family members in bronchial brushings controls (n = 6) and asthma patients (n = 20) were measured by quantitative PCR. **C)** *IL-4Rα* transcript levels in bronchial brushings from asthma patients (n= 38) and healthy controls (n = 17) were measured by quantitative PCR. **D)** Spearman’s rank-order correlation assay between epithelial *IL-4Rα* and *CDH26* transcript levels in asthma patients (n = 38). **E)** Spearman’s rank-order correlation assays between epithelial *CDH26* transcript levels and three-gene-mean of *CLCA1, POSTN* and *SERPINB2* in asthma patients (n=56). **F)** Spearman’s rank-order correlation assays between epithelial *IL-4Rα* transcript levels and three-gene-mean of asthma patients (n=26). **G)** Representative images of CDH26 (green) and IL-4Rα (red) immunofluorescence staining in in bronchial biopsies from controls and asthma patients. Nuclei were stained with DAPI (blue). Scale bar, 50 μm. **H)** Quantification of CDH26 and IL-4Ra immunofluorescent intensity was performed using ImageJ. **I, J)** *MUC5AC* (*I*) and *MUC5B* (*J*) transcript levels in bronchial brushings from asthma patients (n= 38) and healthy controls (n = 17) were measured by quantitative PCR. **K, L)** Spearman’s rank-order correlation assays between epithelial *MUC5AC* (*K*), *MUC5B* (*L*) and *CDH26* transcript levels in asthma patients (n=35). **M, N)** Correlation assays between epithelial *CDH26* transcript levels and percentage of eosinophils in induced sputum (*M*) and FeNO (*N*) of asthma patients (n=56). **O)** *CCL26* transcript levels in bronchial brushings from asthma patients (n=52) and healthy controls (n =13) were measured by quantitative PCR (two-tailed Mann–Whitney test). **P)** Correlation assay between epithelial *CCL26* and *CDH26* transcript levels in asthma patients (n=52). Data are mean ± SD. ***P < 0.001; ****P < 0.0001.

*MUC5AC* transcripts were increased, whereas *MUC5B* transcripts were decreased in bronchial brushings from asthma patients compared to controls (Figure 6I-6J). Epithelial *CDH26* transcript levels were strongly correlated with *MUC5AC* or *MUC5B* expression, supporting that CDH26 promotes aberrant mucin expression in asthma (Figure 6K-6L). Moreover, epithelial *CDH26* transcript levels strongly correlate with the percentage of eosinophils in sputum and FeNO, two parameters reflecting airway eosinophilia (Figure 6M-6N). In addition, eosinophilic chemokine *CCL26* transcripts increased and correlated with *CDH26* expression in asthma patients (Figure 6O-6P). These are consistent with our *in vivo* and *in vitro* findings that CDH26 promotes eosinophilic chemokine expression and airway eosinophilia.

## DISCUSSION

IL-4R signaling pathway plays a critical role in allergic diseases including asthma^6,42^. In our asthmatic patients, IL-4Rα expression in airway epithelium was significantly increased compared to healthy controls, suggesting epithelial IL-4R signaling is upregulated in asthma. It was reported that enhanced STUB1-mediated IL-4Rα degradation might be a feedback mechanism to dampen the upregulated IL-4R signaling in asthma^21^. However, the mechanism underlying epithelial IL-4R upregulation in asthma remains unknown. Herein, we reported that IL-4R ligand upregulates CDH26 in airway epithelial cells, and CDH26 in turn amplifies epithelial IL-4R signaling pathway. This feedforward mechanism leads to, at least in part, the epithelial IL-4R upregulation in asthma patients.

CDH26, an epithelial transmembrane protein belonging to cadherin family, was reported to be upregulated in asthma^28,31^. In our asthma patients, compared to other 10 cadherin family members, CDH26 was the only one that was significantly upregulated in airway epithelium. Interestingly, immunostaining revealed that CDH26 and IL-4Rα proteins were colocalized with each other in airway epithelium of asthma patients and in a mouse model of allergic disease. In support of the amplifying role of CDH26 in epithelial IL-4R signaling, upregulated CDH26 expression paralleled with increased IL-4Rα expression in airway epithelium of asthma patients and the mouse model. However, IL-4Rα expression in airway epithelium was significantly reduced in *Cdh26*^*-/-*^ mice. Type II IL-4R is consisted of IL-4Rα and IL-13Rα1 and is predominantly expressed on airway epithelial cells. IL-13, the ligand of type II IL-4R, increased CDH26 expression as well as IL-4Rα and IL-13Rα1 in our *in vitro* system. Intriguingly, knockdown of CDH26 suppressed IL-13-induced IL-4Rα and IL-13Rα1 expression, and inhibited the downstream signaling including Jak1 and Stat6 phosphorylation. The existence of an amplifying mechanism for IL-4R in hematopoietic cells including macrophages and mast cells has been reported^22^. Here, we reported that CDH26 is an amplifier of IL-4R signaling in non-hematopoietic airway epithelial cells. However, whether CDH26 directly interact with IL-4Rα and the mechanism by which CDH26 regulates IL-4Rα expression require further study.

Airway mucus cell metaplasia and mucus overproduction is one of the hall mark characteristics of asthma, which is strictly dependent on the expression of IL-4Rα and Stat6 in airway epithelium^16,17^. Cdh26 deficiency nearly blocked mucus cell hyperplasia (PAS-positive cells reduced by 86%) and aberrant expression of Muc5ac (Mac5ac-positive cells reduced by 75%) in the mouse model. This suggests that CDH26 plays a critical role in epithelial IL-4R signaling-medicated mucus overproduction in asthma. Consistently, knockdown of CDH26 completely suppressed IL-13 induced MUC5AC upregulation and MUC5B downregulation in HBE cells. Moreover, *CDH26* expression is strongly correlated with *MUC5AC* and *MUC5B* expression in bronchial brushings from asthmatic patients. Interestingly, CDH1 (E-cadherin) was reported to promote MUC5AC production via EGFR-mediated cell differentiation^43^. However, *CDH1* transcript levels were not increased in our asthma patients (Figure 6B).

Airway eosinophilic inflammation is another prominent feature of asthma^1,44^. In our asthmatic patients, *CDH26* expression strongly correlates with sputum eosinophilia and FeNO. However, Cdh26 deficiency only slightly alleviated allergen-induced airway eosinophilia. This may be partly attributed to the reduced expression of eotaxins including Ccl11, Ccl24 and Ccl26 in *Cdh26*^*-/-*^ mice. Additionally, it has been reported that allergen-induced airway eosinophilia is only partially dependent on Stat6 signaling^5^. In cultured HBE cells, we confirmed that CCL11, CCL24, and CCL26 expression were dependent on IL-4R signaling, and CDH26 knockdown suppressed the expression of these chemokines. However, we cannot exclude the possibility that Cdh26 may directly promotes airway eosinophilia because CDH26 is reported to be an epithelial receptor binding integrin α and may play a role in eosinophils recruitment or activation^26^.

There are several limitations of our study. First, using the global *Cdh26* knockout mouse, we cannot exclude the possibility that CDH26 may also affect IL-4R signaling in cell types other than epithelial cells. Second, the mechanism by which CDH26 amplifies epithelial IL-4R signaling requires further investigation. Third, the subjects included in our study were mild-to-moderate asthma, the expression of CDH26 and its association with epithelial IL-4R expression in severe asthma is unclear.

In conclusion, we uncover a novel mechanism underlying IL-4R upregulation in airway epithelial cells in asthma. As an amplifier of epithelial IL-4R signaling, CDH26 represents a potential therapeutic target for airway mucus production in asthma.

## Data Availability

All data referred to in the manuscript are provided in the manuscript and supplements.

## ACKNOWLEDGEMENTS

We thank the participants who volunteered for the study; Xiaoling Rao and Mei Liu for bronchoscopy support; Wang Ni and Shixin Chen for spirometry measurement.

