## Supplementary methods, tables and figures for "CDH26 amplifies airway epithelial IL-4 receptor α signaling in asthma"

### Histology and immunofluorescence staining

For immunofluorescence staining, tissue sections were incubated with anti-CDH26 antibody (Sigma-Aldrich Chemie Gmbh, Munich, Germany, 1:100 dilution) and then incubated with Cy3 donkey anti-rabbit secondary antibody (Boster, Wuhan, China). Double staining was performed in immunofluorescence staining by using two different host species antibodies. The combinations of the primary antibodies include CDH26 (Sigma-Aldrich, 1:100 dilution) with human IL-4Rα antibody (R&D system,1:20 dilution), CDH26 (Sigma-Aldrich, 1:100 dilution) with mouse IL-4Rα antibody (R&D system, 1:200 dilution). Paraffin tissue sections were deparaffinization, rehydration, and processed with pH9.0 Tris-EDTA retrieval buffer (Servicebio, Wuhan, China) before blocking with 10% donkey serum. Paraffin tissue sections were incubated with primary antibodies at 4°C overnight and then incubated with Cy3 or FITC conjugated secondary antibodies (donkey-anti mouse or rabbit IgG, Wuhan, China) at 1:200 in the dark for 1 hour at room temperature. Nuclei were stained with DAPI (Servicebio, Wuhan, China). All photographs were taken by using a fluorescence microscope (Olympus Corporation, Japan).

### Assessment of airway inflammation

Cell counts for macrophages, eosinophils, lymphocytes, and neutrophils in bronchoalveolar lavage fluid (BALF) were performed. The severity of peri-bronchial inflammation in H&E-stained mouse lung sections was scored using the following features: 0, normal; 1, few cells; 2, a ring of inflammatory cells (1 cell layer deep); 3, a ring of inflammatory cells (2–4 cells deep); 4, a ring of inflammatory cells (> 4 cells deep).

**Periodic acid-Schiff (PAS) staining**

Lung sections were stained with PAS (Servicebio, Wuhan, China) for detection of mucus. The number of PAS-staining-positive cells was counted in five random fields for each lung section at 200× magnification.

**The sequence of CDH26 siRNA**

The sequence of CDH26 siRNA (sense strand) was:5’-GGGACUUUCCCAGAA GCAATTUUGCUUCUGGGAAAGUCCCTT-3’. The sequence of negative control siRNA was: sense: 5’-UUCUCCGAACGUACGUTT-3’; antisense: 5’-ACGUG ACACGUUCGGAGAATT-3’.

**SUPPLEMENTARY TABLES**

**Table S1. Subject characteristics.**

| | Healthy controls | Asthma patients | P value |
|---|---|---|---|
| Number | 17 | 56 | |
| Age, y | 35.94+9.43 | 38.09+11.33 | 0.574 |
| Sex, M: F, %F | 5:12, 70.6 | 13:45, 80.4 | 0.604 |
| Body mass index | 22.15+3.32 | 22.33+3.19 | 0.716 |
| $FEV_1$, %predicted | 89.82+32.63 | 79.15+19.89 | 0.007 |
| Methacholine PD20, mg | 2.505+0 | 0.177+0.34 | <0.0001 |
| Sputum eosinophil, % | 1.63+2.77 | 17.03+18.85 | <0.0001 |
| FeNO, ppb | 15.612+11.10 | 100.80+60.84 | <0.0001 |

Note: Values were presented as mean ± SD.

Abbreviations: FeNO, fraction of exhaled nitric oxide; FEV1, forced expiratory volume in the first second; PD20, provocative dosage required to cause a 20% decline in FEV1. The minimal and maximal provocative dosages were 0.01 and 2.505 mg, respectively.

**Table S2. Primers for quantitative PCR**

| Gene | Species | Primer sequence (5'-3') | |
|---|---|---|---|
| GUSB | Human | Forward | GTCTGCGGCATTTTGTCGG |
| | | Reverse | CACACGATGGCATAGGAATGG |
| GAPDH | Human | Forward | AAGGTGAAGGTCGGAGTCAAC |
| | | Reverse | GGGGTCATTGATGGCAACAATA |
| CDH26 | Human | Forward | CCTACCTCACGTCTACAGCGA |
| | | Reverse | TTGAACCCAAAGAGTCCAGCA |
| IL-4Rα | Human | Forward | AAATCGTGAACTTTGTCTCCGT |
| | | Reverse | CCCAGTGCCCTCTACTCTCAT |
| MUC5AC | Human | Forward | CGACAACTACTTCTGCGGTGC |
| | | Reverse | GCACTCATCCTTCCTGTCGTT |
| MUC5B | Human | Forward | CCCGTGTTGTCATCAAGGC |
| | | Reverse | CAGGTCTGGTTGGCGTATTTG |
| CCL26 | Human | Forward | TTGAGGCTGAGCCAAAGACC |
| | | Reverse | GCCCTTCTCAGGTTTCTCCC |
| CCL24 | Human | Forward | ACATCATCCCTACGGGCTCT |
| | | Reverse | CTTGGGGTCGCCACAGAAC |
| CCL11 | Human | Forward | GAAAGCTGTGATCTTCAAGACC |
| | | Reverse | GGCTTTGGAGTTGGAGATTTTT |
| Gapdh | Mouse | Forward | AGAGAGGCCCAGCTACTCG |
| | | Reverse | GGCACTGCACAAGAAGATGC |
| Cdh26 | Mouse | Forward | CCAGCAGAAATCCTTCCAAGGAG |
| | | Reverse | CGATCCAAGAGAAGATGGGTTCTC |
| Il-4Rα | Mouse | Forward | TGACCTCACAGGAACCCAGGC |
| | | Reverse | GAACAGGCAAAACAACGGGAT |
| Muc5ac | Mouse | Forward | CAGGACTCTCTGAAATCGTACCA |
| | | Reverse | GAAGGCTCGTACCACAGGG |
| Muc5b | Mouse | Forward | CTACTCGAACTGCTTGTTTGAC |
| | | Reverse | CTTGTAGCAAACTTTGTCCCTC |
| Ccl26 | Mouse | Forward | ATCCCATGGAGCTGGGTGTA |
| | | Reverse | CCTGGCTGGACACAGAATTG |
| Ccl24 | Mouse | Forward | CAGCCTTCTAAAGGGGCCAA |
| | | Reverse | GCTGGTCTGTCAAACCCCAA |
| Ccl11 | Mouse | Forward | CAACAGATGCACCCTGAAAGC |
| | | Reverse | TGATATTCCCTCAGAGCACGTCTT |
| Il-4 | Mouse | Forward | GTCATCCTGCTCTTCTTTCTCG |
| | | Reverse | CTCTCTGTGGTGTTCTTCGTTG |
| Il-13 | Mouse | Forward | CTGAGCAACATCACACAAGACC |
| | | Reverse | AATCCAGGGCTACACAGAACC |

**SUPPLEMENTARY FIGURES**

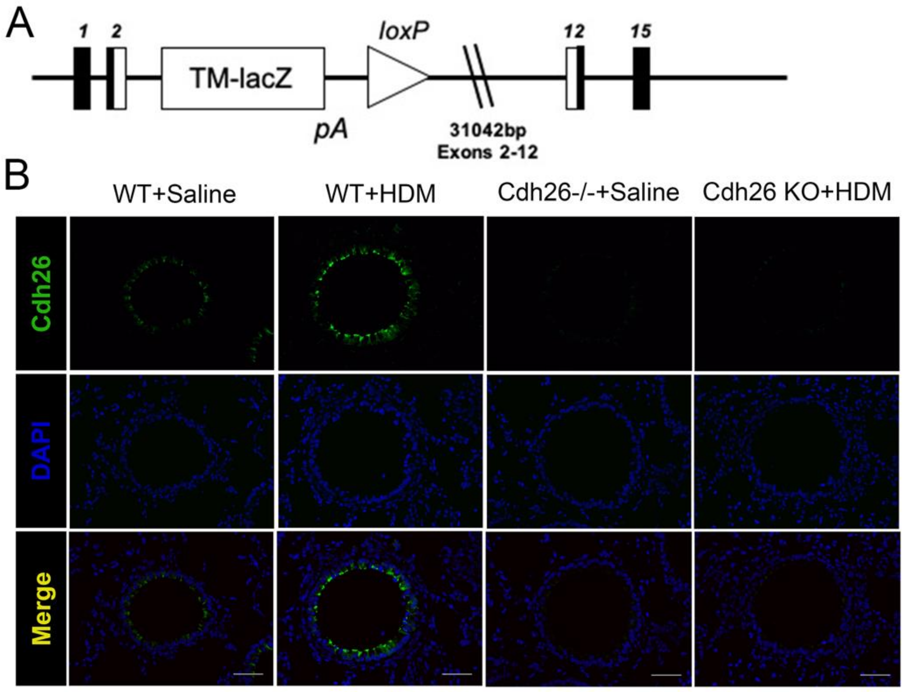


**Figure S1. Generation of *Cdh26$^{-/-}$* mice. A)** Primer Strategy and scheme of *Cdh26$^{-/-}$* mice. Exon 2 to exon 12 of cdh26 gene including 31,042 base pair was knocked out. **B)** Representative images of Cdh26 immunofluorescence staining in mouse lung sections. Scale bar, 50 μm. n =5-8 mice per group.

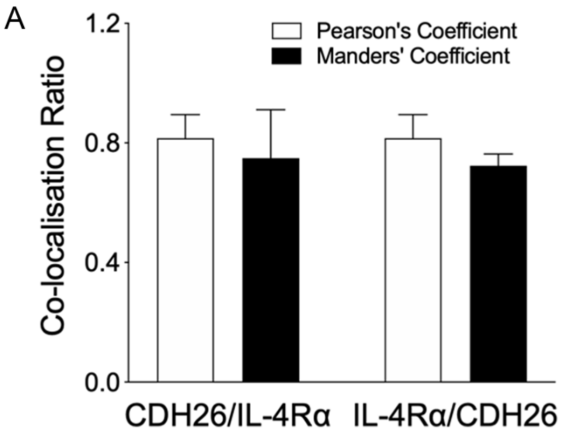


**Figure S2. Cdh26 and Il-4R are co-located in airway epithelium in mouse.**

Quantitative analysis of protein colocalization between Cdh26 and Il-4Rα proteins by Pearson and Manders' coefficients. Colocalization of Cdh26 and Il-4Rα were analyzed using ImageJ plug-in Coloc2. Data are mean ± SD.

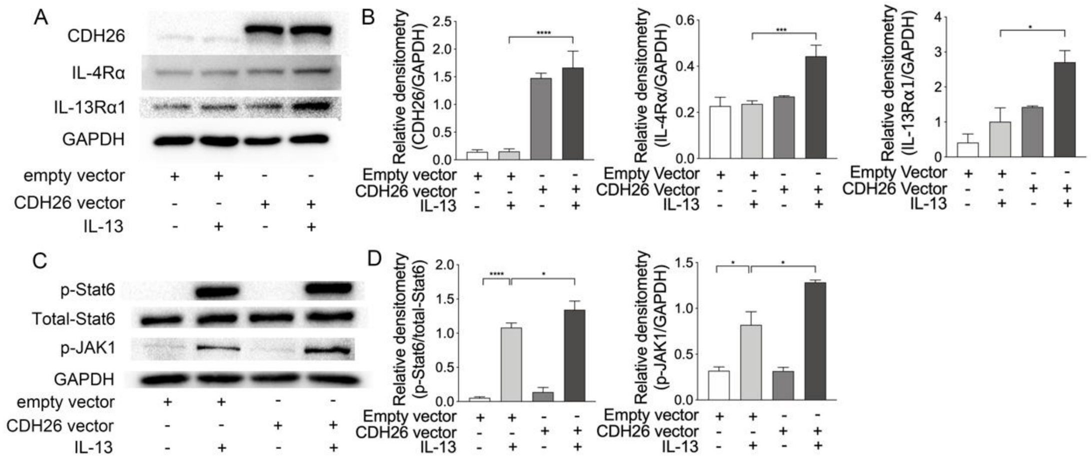

**Figure S3. CDH26 promotes IL-13-induced IL-4Rα expression, JAK-1 and Stat6 phosphorylation. A)** The protein levels of IL-4Rα, IL-13Rα1 in BEAS-2B cells after transfection with empty or CDH26 cDNA expression vector with or without IL-13 stimulation at 48h were detected by Western blotting. **B)** Densitometry assay of the Western blotting results was analyzed using ImageJ. **C)** IL-13 stimulation increased JAK1 and STAT6 phosphorylation at 30min, and CDH26 vector transfection contributed IL-13-induced JAK1 and STAT6 phosphorylation. Densitometry of phospho-JAK1 relative to total GAPDH and phosphor-STAT6 relative to total STAT6. **D)** Densitometry assay of the Western blotting results was analyzed using ImageJ. The data are representative of three independent experiments. Data are mean ± SD. *P < 0.05; **P < 0.01; ***P < 0.001; ****P < 0.0001(one-way ANOVA followed by Tukey's multiple comparison test).

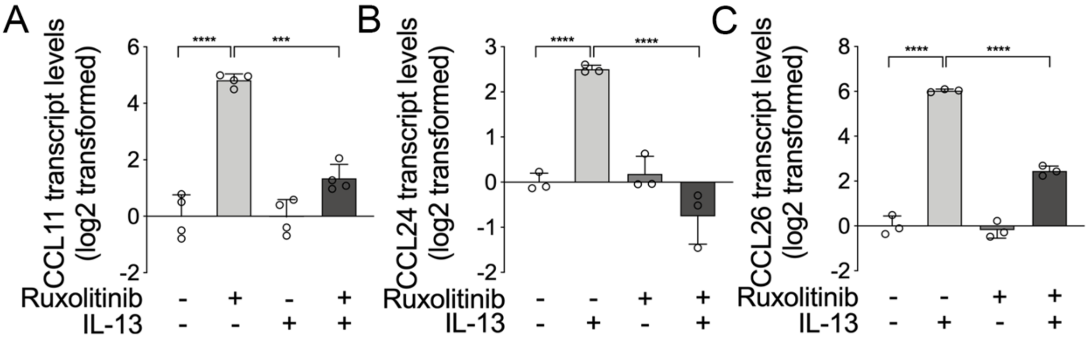


**Figure S4. Selective JAK1/2 inhibitor Ruxolitinib blocked IL-13-induced CCL11, CCL24, CCL26 transcript expression. A-C)** BEAS-2B cells were treated with vehicle or ruxolitinib (100nM) with or without IL-13 (20ng/mL) stimulation for 48h. The mRNA levels of *CCL11* (*A*), *CCL24* (*B*), *CCL26* (*C*) in BEAS-2B cells were determined by quantitative PCR. The transcript level was expressed as log2 transformed and relative to the mean of control group. n = 3-4 wells per group. Data are mean ± SD. *P < 0.05; **P < 0.01; ***P < 0.001; ****P < 0.0001(one-way ANOVA followed by Tukey's multiple comparison test).

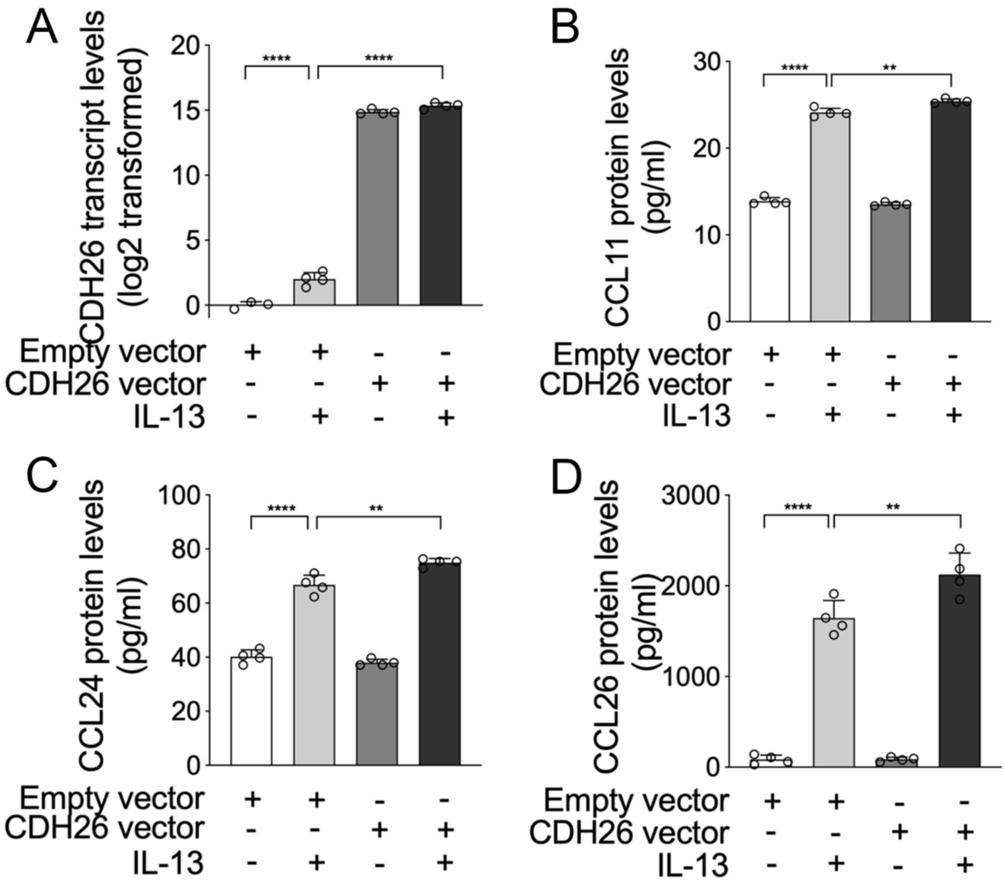


**Figure S5. CDH26 regulates Eotaxin-1, Eotaxin-2, Eotaxin-3 expression in human bronchial epithelial cells. A)** The transcript levels of *CDH26* after transfection with empty or CDH26 cDNA expression vector with or without IL-13 (20ng/mL) stimulation for 48h were detected by quantitative PCR. B-D) The protein levels of CCL11 (*B*), CCL24 (*C*) and CCL26 (*D*) in cell culture media after transfection empty or CDH26 cDNA expression vector with or without IL-13 stimulation were detected by ELISA. n = 4 wells per group. Data are mean ± SD. *P < 0.05; **P < 0.01; ***P < 0.001; ****P < 0.0001(one-way ANOVA followed by Tukey's multiple comparison test).
